# Factors Associated with Malaria Vaccine Hesitancy Among Caregivers of Children 6-59 Months, In Ugenya Sub County, Siaya County, Kenya

**DOI:** 10.64898/2026.04.21.26351425

**Authors:** Jane Muita, Tom Olewe, Evitar Aluoch Ochieng’

## Abstract

**Background:** Malaria remains a leading public health burden in sub-Saharan Africa, disproportionately affecting children under five years. In response, Kenya introduced the RTS,S/AS01 malaria vaccine in selected regions, including Siaya County where malaria transmission is endemic. Despite this milestone, uptake has been inconsistent, with hesitancy emerging as a significant barrier.

**Objective:** This study aimed to determine factors associated with malaria vaccine hesitancy among caregivers of children 6-59 months in Ugenya Subcounty, Siaya County.

**Methodology:** A cross-sectional mixed methods design was employed involving 425 caregivers and 15 healthcare workers and County health officials between January to February 2025. Quantitative data were collected using structured questionnaires and analyzed in Stata version 17 through descriptive statistics, bivariate analysis at 20% significance threshold, and multivariable logistic regression at 5% level to determine key factors associated with malaria vaccine hesitancy. Qualitative data from 15 key informant interviews were transcribed verbatim and thematically analyzed using NVivo. Thematic analysis, guided by a predefined codebook, was used to identify recurring patterns and extract key themes, which were illustrated with direct quotations from participants

**Results:** Overall, 42.9% of caregivers (n=181; 95% CI: 38.9%–47.3%) reported hesitancy. Significant predictors included caregiver age, marital status, family size, access to health facilities, and vaccine availability. Single caregivers, those from smaller households, and those facing health facility access challenges were more likely to be hesitant to malaria vaccine. Despite high levels of knowledge, misconceptions and misinformation about vaccine safety, often spread via social media persisted. Conversely, caregivers relying on healthcare workers and mainstream media showed greater acceptance of malaria vaccine.

**Conclusion and Recommendations:** Malaria vaccine hesitancy remains significant at **42.9%**, driven by demographic factors such as younger age, single status, and smaller household size. Structural barriers including limited vaccine availability and poor access to health facilities further contribute to reluctance. Although knowledge and awareness were high, misinformation, particularly from social media, persisted, while information from healthcare workers improved acceptance. Addressing these gaps through targeted community engagement, improved access, and trusted communication channels is essential to increase uptake of malaria vaccine.

## CHAPTER ONE: INTRODUCTION

### 1.0 Background

Malaria remains one of the world’s most persistent and deadly infectious diseases, posing a significant public health threat globally. Despite being both preventable and treatable, the disease continues to claim hundreds of thousands of lives each year. According to the Centers for Disease Control and Prevention (CDC) and World Health Organization (WHO), nearly half of the global population is at risk of malaria, with an estimated 249 million cases recorded worldwide in 2022 (CDC, 2022; WHO, 2023). A sharp increase from 233 million cases was reported in 2019 (WHO, 2023). Tragically, 627,000 people died from malaria in 2022, with 481,000 of those deaths occurring among children under the age of five. Approximately 96% of all malaria-related deaths took place in sub-Saharan Africa, highlighting the region’s disproportionate burden (WHO, 2023). Africa continues to bear the brunt of malaria’s devastating impact, particularly among vulnerable groups such as young children and pregnant women. Every minute, a child succumbs to malaria, and one in three pregnant women experiences severe illness due to the infection (WHO, 2018). In 2021, UNICEF reported that 80% of all malaria-related deaths in Africa were among children under five (UNICEF, 2021). Additionally, an estimated 819,000 low-birth-weight babies were born due to malaria in pregnancy, further compounding the public health crisis (CDC, 2021). The economic toll is equally staggering. Despite an increase in global malaria funding from $3.5 billion in 2021 to $4.1 billion in 2022, this still falls short of the $7.8 billion target required to meet the Global Technical Strategy goals (UNICEF, 2024). Years of underinvestment, rising insecticide and drug resistance, and disruptions caused by emergencies like the COVID-19 pandemic have weakened malaria control efforts.

Kenya, like many African nations, faces significant challenges in malaria control. While WHO promotes comprehensive strategies such as indoor residual spraying, insecticide-treated nets, larval source management, and timely treatment, their implementation is hindered by climate change, inadequate health infrastructure, and population mobility (Solomon *et al*., 2019). These barriers have led public health stakeholders to explore innovative approaches, including vaccination.

Vaccination is recognized as one of the most successful public health interventions globally, credited with the eradication of smallpox and progress in controlling diseases like polio (Birkett, 2010). In 2017, WHO initiated pilot testing of the RTS,S/AS01 (Mosquirix) malaria vaccine in Kenya, Malawi, and Ghana. Following promising results such as a 30% reduction in severe malaria cases and a 21% drop in hospital admissions; WHO officially recommended its wider use in October 2021. The vaccine, administered in four doses between 6 and 24 months of age, targets the pre-erythrocytic stage of *Plasmodium falciparum* and has demonstrated 39% efficacy against clinical malaria (WHO, 2017; WHO, 2018).

In 2023, WHO approved a second vaccine, R21/Matrix-M, developed by the University of Oxford. In trials conducted in Burkina Faso, R21 demonstrated 77% efficacy over a 12-month follow-up, marking a significant advancement in malaria vaccine development, (WHO, 2023). Despite these scientific breakthroughs, malaria vaccines uptake level remains lower than expected especially in the lower– and middle-income countries (LMIC), in part due to growing vaccine hesitancy, (Sulaiman *et al*., 2025).

Vaccine hesitancy is defined as the delay in acceptance or refusal of vaccines despite their availability. Vaccine hesitancy is an emergent challenge in vaccine rollout, (MacDonald, 2015). Studies show that even among populations with access to immunization services, hesitancy persists due to personal beliefs, cultural influences, misinformation, mistrust in health systems, and broader social and political dynamics (Busby *et al*., 2015). Addressing these complex and context-specific barriers is essential to improving the uptake of malaria vaccines and ensuring their life-saving potential reaches the most vulnerable.

## MATERIAL AND METHODS

### 3.0 Study Design, Area and Population

This was a population based analytical cross-sectional study of malaria vaccine hesitancy among caregivers of children 6-59 months, healthcare workers and Siaya County health officials. This study employed a mixed study design. The study was conducted in Siaya County, Ugenya sub county in Kenya in the study was carried out in the community among caregivers of children 6-59 months, among healthcare providers, and county health officials of Siaya County.

### 3.1 Sample Size Determination

The sample size was calculated using Fischer formula, (Egbuchulem, 2023) of;

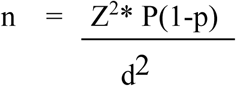

Where:

n= required sample size,

Z= Z-score for 95% Confidence Level (1.96)

P= Estimated proportion of uptake (0.48)

d= Marginal error (0.05)

Therefore:

n = 1.96*1.96*0.48 (1-0.48)/0.05*0.05 =386.4

The study’s sample size was modified to 425 caregivers to account for lack response.

## Results and Discussion

### Socio Demographic and Economic Factors Associated with Malaria Vaccine Hesitancy

#### Socio Demographic and Economic Characteristics of the Study Participants

A total of four hundred and twenty-five (425) caregivers were invited to participate in the study, all of whom completed the survey, resulting in a 100% response rate. The participants’ ages ranged from 15 to 99 years, with the majority falling between 26-35 years old (35.8%). The average age was 35 years. A large proportion of the participants were females (87.1%, n=370), while males accounted for 13% (n=55). Most caregivers were the children’s biological parents (89.2%, n=379), whereas 10.8% (n=46) were identified as the children’s guardians.

Among the participants, 98.6% (n=419) identified as Christians, 0.7% (n=3) as Muslims and 0.7% (n=3) as traditionalist/herbalist. Most of the caregivers, 63.1% (n=268) reported having a family size of <4 members. Regarding marital status, 77.9% (n=331) of the caregivers were married, 13.4% (n=57) single and 7.1% (n=30) widowed. Overall, income levels were generally low among the respondents. A majority, 63.1%, (n=268) reported a monthly income below Ksh 10,000, while only 14.8% (n=63) earned more than Ksh 15,000.

In terms of employment status, 72.5% (n=308) of caregivers were unemployed. A substantial proportion of the caregivers, 74.8% (n=318) were either unemployed, students, or retired, 15.8% (n=67) were self-employed, and 9.4% (n=40) were formally employed. Regarding education levels, 58.6% (n=249) of caregivers had attained primary education, 22.6% (n=96) had attained secondary school, 8.2% (n=35) had tertiary education, and 10.6% (n=45) had no formal education. The caregivers resided in three electoral wards: with 38.2% (n=162) North Ugenya, 33.7% (n=143) West Ugenya and 28.2% (n=120) East Ugenya.

**Table 1:**
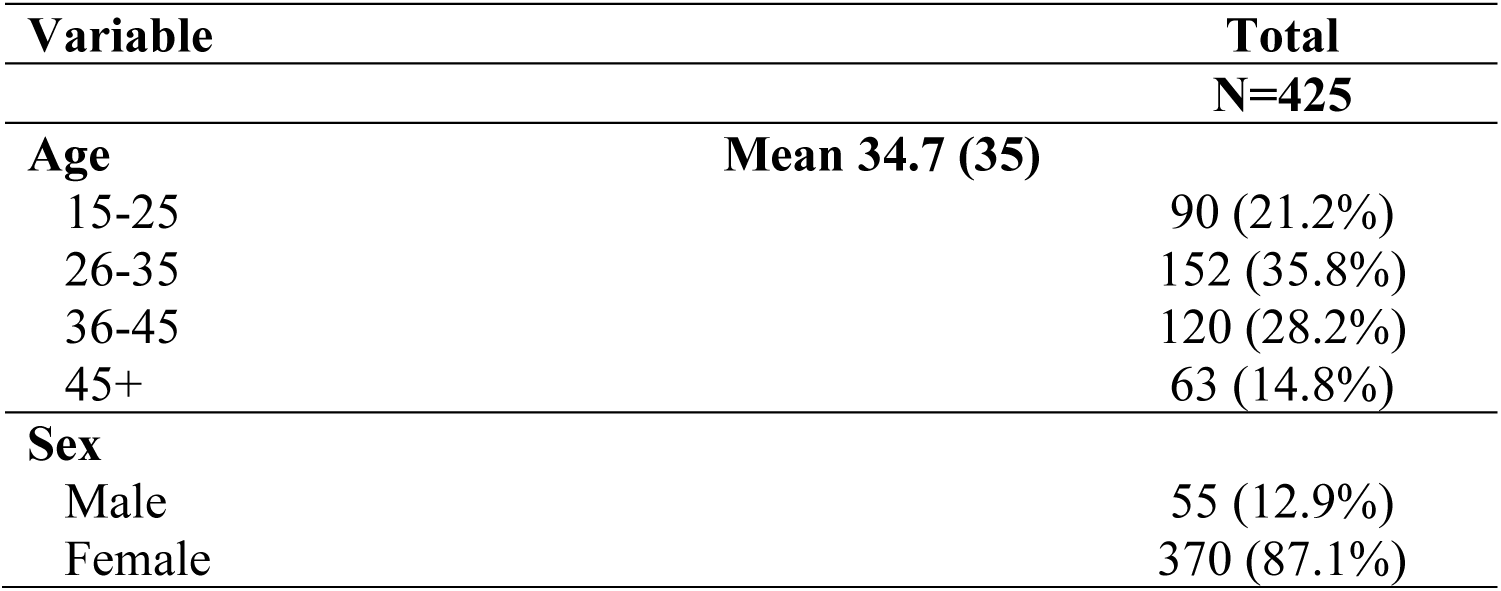
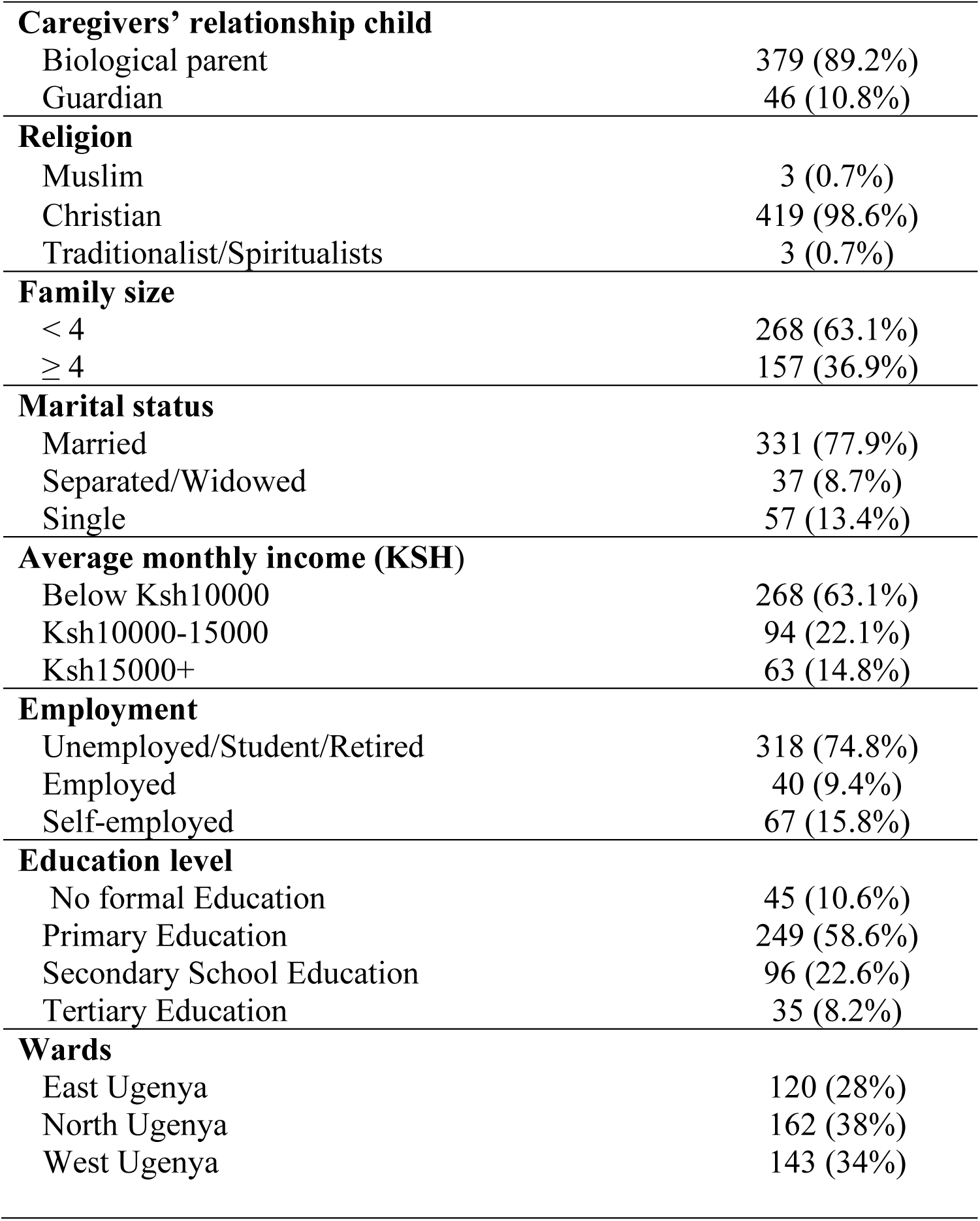
Univariate analysis of socio-demographic and economic characteristics of caregivers of children under 5yrs in Ugenya sub county, Siaya County, (N = 425)

| Variable | Total<br>N=425 |
| --- | --- |
| **Age** | **Mean 34.7 (35)** |
| 15-25 | 90 (21.2%) |
| 26-35 | 152 (35.8%) |
| 36-45 | 120 (28.2%) |
| 45+ | 63 (14.8%) |
| **Sex** |  |
| Male | 55 (12.9%) |
| Female | 370 (87.1%) |

|  |  |  |
| --- | --- | --- |
| **Caregivers' relationship child** |  |  |
| Biological parent |  | 379 (89.2%) |
| Guardian |  | 46 (10.8%) |
| **Religion** |  |  |
| Muslim |  | 3 (0.7%) |
| Christian |  | 419 (98.6%) |
| Traditionalist/Spiritualists |  | 3 (0.7%) |
| **Family size** |  |  |
| < 4 |  | 268 (63.1%) |
| ≥ 4 |  | 157 (36.9%) |
| **Marital status** |  |  |
| Married |  | 331 (77.9%) |
| Separated/Widowed |  | 37 (8.7%) |
| Single |  | 57 (13.4%) |
| **Average monthly income (KSH)** |  |  |
| Below Ksh10000 |  | 268 (63.1%) |
| Ksh10000-15000 |  | 94 (22.1%) |
| Ksh15000+ |  | 63 (14.8%) |
| **Employment** |  |  |
| Unemployed/Student/Retired |  | 318 (74.8%) |
| Employed |  | 40 (9.4%) |
| Self-employed |  | 67 (15.8%) |
| **Education level** |  |  |
| No formal Education |  | 45 (10.6%) |
| Primary Education |  | 249 (58.6%) |
| Secondary School Education |  | 96 (22.6%) |
| Tertiary Education |  | 35 (8.2%) |
| **Wards** |  |  |
| East Ugenya |  | 120 (28%) |
| North Ugenya |  | 162 (38%) |
| West Ugenya |  | 143 (34%) |

#### 4.0.1 Bivariate analysis of Socio Demographic and Economic Characteristics

A bivariate analysis of the socio demographic and economic characteristics (**Table 2**) was conducted to examine the association with malaria vaccine hesitancy at a 20% significance level. *Age* was found to be significantly associated with malaria vaccine hesitancy (p = 0.013). The highest proportion of hesitancy was observed among caregivers aged 36–45 years (52%), while those aged 26–35 and above 45 years and above reported lower levels of malaria vaccine hesitancy at 35% each.

**Table 2:** Association between socio-demographic and economic characteristics of the Caregivers of children 6-59 months and Malaria vaccine hesitancy, in Ugenya subcounty, Siaya County.

|  | No<br>N=244 | Yes<br>N=181 | p-value |
| --- | --- | --- | --- |
| Age |  |  |  |
| 15-25 | 46 (51%) | 44 (49%) | 0.013 |
| 26-35 | 99 (65%) | 53 (35%) |  |
| 36-45 | 58 (48%) | 62 (52%) |  |
| 45+ | 41 (65%) | 22 (35%) |  |
| Sex |  |  |  |
| Male | 29 (53%) | 26 (47%) | 0.45 |
| Female | 215 (58%) | 155 (42%) |  |
| Caregivers Relationship Child |  |  |  |
| Biological parent | 210 (55%) | 169 (45%) | 0.017 |
| Guardian | 34 (74%) | 12 (26%) |  |
| Religion |  |  |  |
| Muslim | 3 (100%) | 0 (0%) | 0.23 |
| Christian | 240 (57%) | 179 (43%) |  |
| Traditionalist/Spiritualists | 1 (33%) | 2 (67%) |  |
| Family size |  |  | <0.001 |
| < 4 | 129 (48%) | 139 (52%) |  |
| >=4 | 115 (73%) | 42 (27%) |  |
| Marital status |  |  |  |
| Married | 196 (59%) | 135 (41%) | 0.012 |
| Separated/Widowed | 25 (68%) | 12 (32%) |  |
| Single | 23 (40%) | 34 (60%) |  |
| Average monthly income (KSH) |  |  |  |
| Below Ksh10000 | 162 (60%) | 106 (40%) | 0.062 |
| Ksh10000-15000 | 44 (47%) | 50 (53%) |  |
| Ksh15000+ | 38 (60%) | 25 (40%) |  |
| Employment |  |  |  |
| Unemployed/Student/Retired | 182 (57%) | 136 (43%) | 0.94 |
| Employed | 24 (60%) | 16 (40%) |  |
| Self-employed | 38 (57%) | 29 (43%) |  |
| Highest Education level |  |  |  |
| No formal Education | 21 (47%) | 24 (53%) | 0.097 |
| Primary Education | 150 (60%) | 99 (40%) |  |
| Secondary School Education | 49 (51%) | 47 (49%) |  |
| Tertiary Education | 24 (69%) | 11 (31%) |  |
| Sub County Ward |  |  |  |
| East Ugenya | 64 (53%) | 56 (47%) | <0.001 |
| North Ugenya | 73 (45%) | 89 (55%) |  |
| West Ugenya | 107 (75%) | 36 (25%) |  |

*Sex* was not significantly associated with malaria vaccine hesitancy (p = 0.45), indicating no difference in hesitancy levels between males (47%) and females (42%). However, a statistically significant association was observed between *caregiver’s relationship to the child* and malaria vaccine hesitancy (p = 0.017). Biological parents of the children reported higher malaria vaccine hesitancy (44.6%) compared to guardians (26%). Religion was not significantly associated with malaria vaccine hesitancy (p = 0.23). Muslim reported no malaria vaccine hesitancy (100%), while Christians and Traditionalists/Spiritualists reported 43% and 67% malaria vaccine hesitancy respectively.

Qualitative analysis revealed that certain religious groups discouraged their members from receiving the malaria vaccine. A Community Health Promoter (CHP) noted that some churches explicitly advised their members against seeking healthcare services, such as vaccinations, as quoted: “**R:** *In the past there used to be some churches that were discouraging congregants from seeking services from the Health Care Facilities (HCF)*.” **KII01_MVH_CHP**.

Additionally, a nurse practitioners reported that some caregivers claimed that syringes were incompatible with their religions as supported by quotation: *Yeah, there are some. They tell you; “Syringes don’t work well with our Religion”, one mama told us. Yeah, there is. It’s called Msambwo. You know, where we are here, we have borders with Luhya’s. So dini ya Msambwo for the Luhya’s, then Luo, we have the Roho.”* **KII02_MVH_NP**

Family size was significantly associated with malaria vaccine hesitancy (*p < 0.001*). Caregivers with smaller family size (<4 members) had higher malaria vaccine hesitancy 52% (n=139) compared to those with larger sizes (≥4 members), who reported lower malaria vaccine hesitancy 27% (n=42). Marital status was also significantly associated with malaria vaccine hesitancy (*p = 0.012*); Unmarried (Single) caregivers had the highest rate of malaria vaccine hesitancy 60% (n=34), followed by married caregivers 41% (n=135) and separated, widowed or divorced 32% (n=12). Average monthly income of the caregivers did had an association with vaccine hesitancy (p = 0.062). Caregivers with monthly income of Ksh 10,000 – 15,000 reported highest malaria vaccine hesitancy 53% (n=50), compared to those earning below 10,000 40% (n=106) above 15,000 40 (n=25) per month.

Employment status of the caregivers was not significantly associated with malaria vaccine hesitancy (p = 0.94). Malaria vaccine hesitancy was reported among 43% (n = 136) of respondents who were unemployed, students, or retired. Similarly, 40.0% (n = 16) of employed caregivers and 43% (n = 29) of self-employed caregivers showed hesitancy towards malaria vaccine, indicating relatively similar proportions across employment categories. This was corroborated by a key informant who reported that “*They don’t have fare and walking to the facility is not easy while carrying the baby.”* **KII08_MVH_CHP”**

Education level was significantly associated with malaria vaccine hesitancy (p = 0.097. Caregivers with no formal education reported highest level of malaria vaccine hesitancy at 53% (n = 24), followed by those with secondary education at 49% (n = 47). Hesitancy was lower among caregivers with primary education at 40% (n = 99), and lowest among those with tertiary education at 31% (n = 11).

The ward of residency was significantly associated with malaria vaccine hesitancy (p < 0.001). Caregivers from North Ugenya reported the highest level of malaria vaccine hesitancy 55% (n=89), compared to those from East Ugenya 47% (n=56), and West Ugenya 25% (n=36).

### 4.1 Individual and psychosocial factors Associated with Malaria Vaccine Hesitancy

#### 4.1.1 Individual and psychosocial characteristics of caregivers of children 6-59 months in Ugenya Subcounty, Siaya County

The study assessed individual and psychosocial factors such as the caregivers’ knowledge, source of information, awareness and willingness to vaccinate their children against malaria (**Table 3**). A majority 91% (n=387) of the caregivers reported good knowledge of malaria vaccine.

**Table 3:** Individual and psychosocial Characteristics of caregivers of children 6-59 months old in Ugenya Subcounty, Siaya County.

|  | **Total**<br>**N=425** |
| --- | --- |
| **Awareness** |  |
| Heard about malaria vaccine |  |
| No | 9 (2%) |
| Yes | 416 (98%) |
| **Source of information** |  |
| Social media | 230 (54%) |
| Community leaders | 238 (56%) |
| Healthcare workers | 314 (74%) |
| Community health partners | 254 (60%) |
| Word of mouth (friends/family/neighbors) | 125 (29%) |
| Mainstream media (radio/tv) | 367 (86%) |
| Print media (newspapers) | 259 (61%) |
| Schools/educational or religious institutions | 38 (9%) |
| **Knowledge** |  |
| Malaria vaccine schedule |  |
| No | 28 (7%) |
| Yes | 397 (93%) |
| Number of malaria vaccine doses |  |
| No | 34 (8%) |
| Yes | 391 (92%) |
| Overall knowledge |  |
| Poor | 38 (9%) |
| Good | 387 (91%) |
| **Negative perception of malaria vaccine** |  |
| It has potential side effect | 30 (7%) |
| I'm worried about the vaccine safety | 28 (7%) |
| **Willingness** |  |
| Has your child been vaccinated |  |
| No | 10 (2%) |
| Yes | 415 (98%) |
| a) Was the decision made willingly |  |
| No | 3 (1%) |
| Yes | 412 (99%) |
| b) If No who made the decision |  |
| Healthcare Worker | 3 (100%) |
| Are you willing to vaccinate |  |
| No | 4 (40%) |
| Yes | 6 (60%) |
| Reasons for unwillingness |  |
| Fear of the side effects | 2 (50%) |
| Lack of awareness about the vaccine | 2 (50%) |

A large majority of caregivers, 98% (n=416) were aware of malaria vaccine. The primary sources of information included mainstream media 88.2% (n=357), healthcare workers 75.5%, (n=314), and community leaders 57.2% (n=238). Other notable sources of malaria vaccine were print media, 62.3% (n=259), social media 55.3% (n=230), and community health partners 61.1% (n=254), showing a blend of traditional and digital platforms being used to reach the public. Less commonly reported sources included word-of-mouth and schools or religious institutions 30.1% (n=125) and 9.1% e(n=38) respectively.

A high proportion 97.7% (n=415) of caregivers reported that their children had received the malaria vaccine. Among those whose children had been vaccinated, 99.3% (n=412) stated that the decision was arrived at willingly. The remaining 0.7% (n=3) revealed that the decision was made by the healthcare worker rather than themselves. Regarding perceived risks of malaria vaccine, 7% (n=30) of the caregivers expressed concerns about the potential side effects, and a similar proportion were concerned about malaria vaccine safety. A nurse practitioner reported that some of the caregivers believed that vaccination could cause paralysis as illustrated in the following quote: *“They believe that if the baby gets immunized, they will be paralyzed. We have a certain village down in Roslyn and it’s almost like, it’s not only malaria, but also all the vaccines. Their babies are not getting immunized.”* **KII03_MVH_NP**

Out of the caregivers surveyed 2.4%, (n=10) reported that their children had not received malaria vaccine. Of these, 60% (n=6) reported that they were willing get the children vaccinated. Among the 40% (n = 4) of respondents who were unwilling to vaccinate, 50%, (n=2) cited fear of the side effects and 50% (n=2) reported lack of awareness about the malaria vaccine as the reasons to their unwillingness as represented in result **Table 3**.

In the qualitative analysis, a key informant reported that despite referrals and home visits, some of the caregivers remained unwilling to take their children for the malaria vaccinations illustrated in the following quote.

“Because even if I do home visits and do a referral some will still not go, and you can’t beat them they’re adults and the baby is also hers.” **KII08_MVH_CHP**

#### 4.1.2 Association between individual and psychosocial factors; and Malaria vaccine hesitancy

The bivariate analysis of individual and psychosocial factors at 20% significance level (α = 0.20), showed that awareness, source of information and knowledge were associated with malaria vaccine hesitancy as shown on **Table 4**.

**Table 4:**
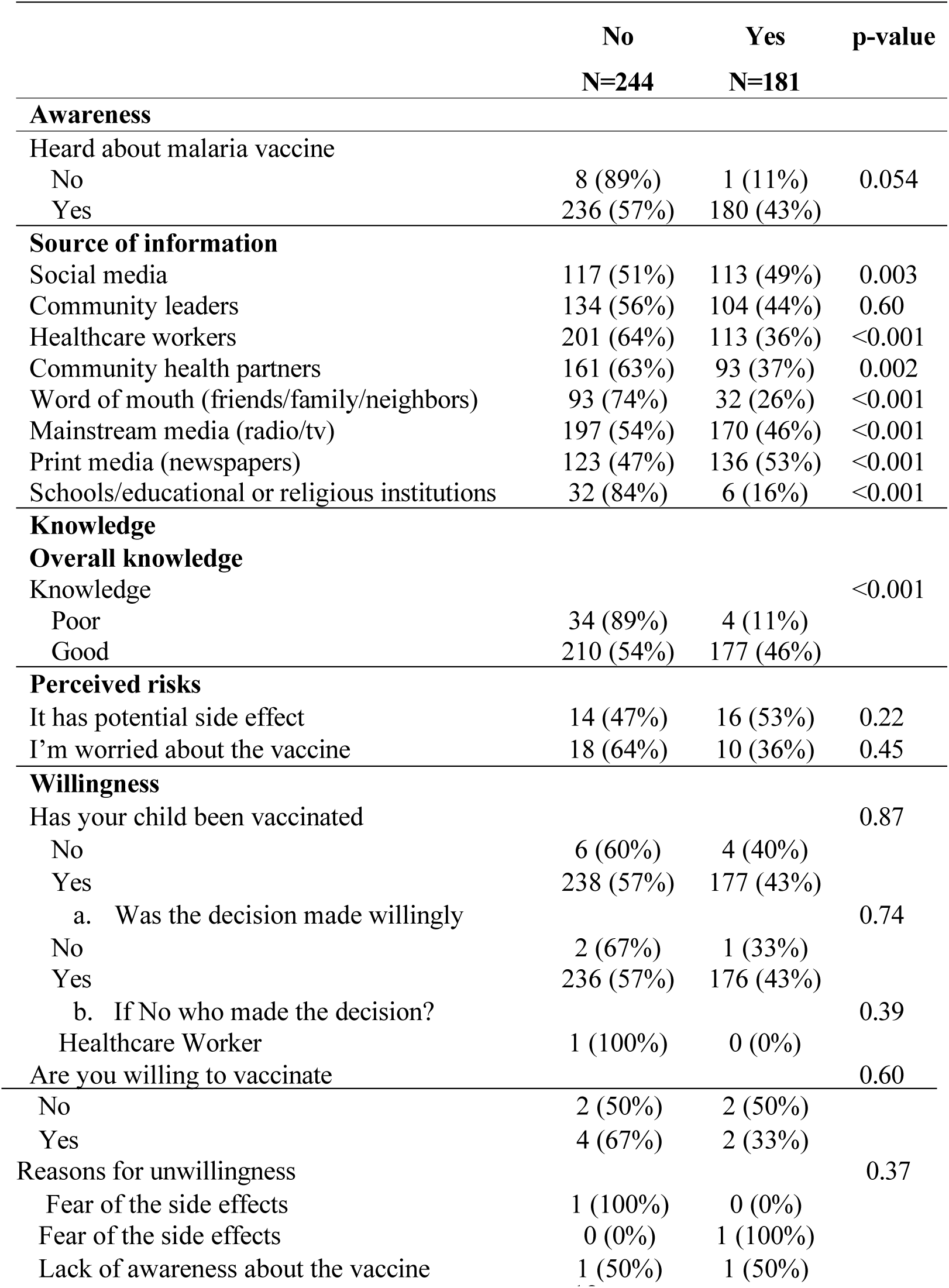
Association between individual and psychosocial characteristics of the Caregiver of children 6-59 months and Malaria vaccine hesitancy, in Ugenya Subcounty, Siaya County.

|  | No<br>N=244 | Yes<br>N=181 | p-value |
| --- | --- | --- | --- |
| **Awareness** |  |  |  |
| Heard about malaria vaccine |  |  |  |
| No | 8 (89%) | 1 (11%) | 0.054 |
| Yes | 236 (57%) | 180 (43%) |  |
| **Source of information** |  |  |  |
| Social media | 117 (51%) | 113 (49%) | 0.003 |
| Community leaders | 134 (56%) | 104 (44%) | 0.60 |
| Healthcare workers | 201 (64%) | 113 (36%) | <0.001 |
| Community health partners | 161 (63%) | 93 (37%) | 0.002 |
| Word of mouth (friends/family/neighbors) | 93 (74%) | 32 (26%) | <0.001 |
| Mainstream media (radio/tv) | 197 (54%) | 170 (46%) | <0.001 |
| Print media (newspapers) | 123 (47%) | 136 (53%) | <0.001 |
| Schools/educational or religious institutions | 32 (84%) | 6 (16%) | <0.001 |
| **Knowledge** |  |  |  |
| **Overall knowledge** |  |  |  |
| Knowledge |  |  | <0.001 |
| Poor | 34 (89%) | 4 (11%) |  |
| Good | 210 (54%) | 177 (46%) |  |
| **Perceived risks** |  |  |  |
| It has potential side effect | 14 (47%) | 16 (53%) | 0.22 |
| I'm worried about the vaccine | 18 (64%) | 10 (36%) | 0.45 |
| **Willingness** |  |  |  |
| Has your child been vaccinated |  |  | 0.87 |
| No | 6 (60%) | 4 (40%) |  |
| Yes | 238 (57%) | 177 (43%) |  |
| a. Was the decision made willingly |  |  | 0.74 |
| No | 2 (67%) | 1 (33%) |  |
| Yes | 236 (57%) | 176 (43%) |  |
| b. If No who made the decision? |  |  | 0.39 |
| Healthcare Worker | 1 (100%) | 0 (0%) |  |
| Are you willing to vaccinate |  |  | 0.60 |
| No | 2 (50%) | 2 (50%) |  |
| Yes | 4 (67%) | 2 (33%) |  |
| Reasons for unwillingness |  |  | 0.37 |
| Fear of the side effects | 1 (100%) | 0 (0%) |  |
| Fear of the side effects | 0 (0%) | 1 (100%) |  |
| Lack of awareness about the vaccine | 1 (50%) | 1 (50%) |  |

There was a significant association between awareness and malaria vaccine hesitancy, (p= 0.054). Caregivers who had heard about malaria vaccine reported higher hesitancy to malaria vaccine (n=180). Knowledge of malaria vaccine was significantly associated with mala hesitancy (p<0.001). Caregivers with good knowledge of the malaria vaccine reported higher levels of hesitancy (46%) compared to those with poor knowledge (11%) as shown in **Table 5**.

**Table 5:**
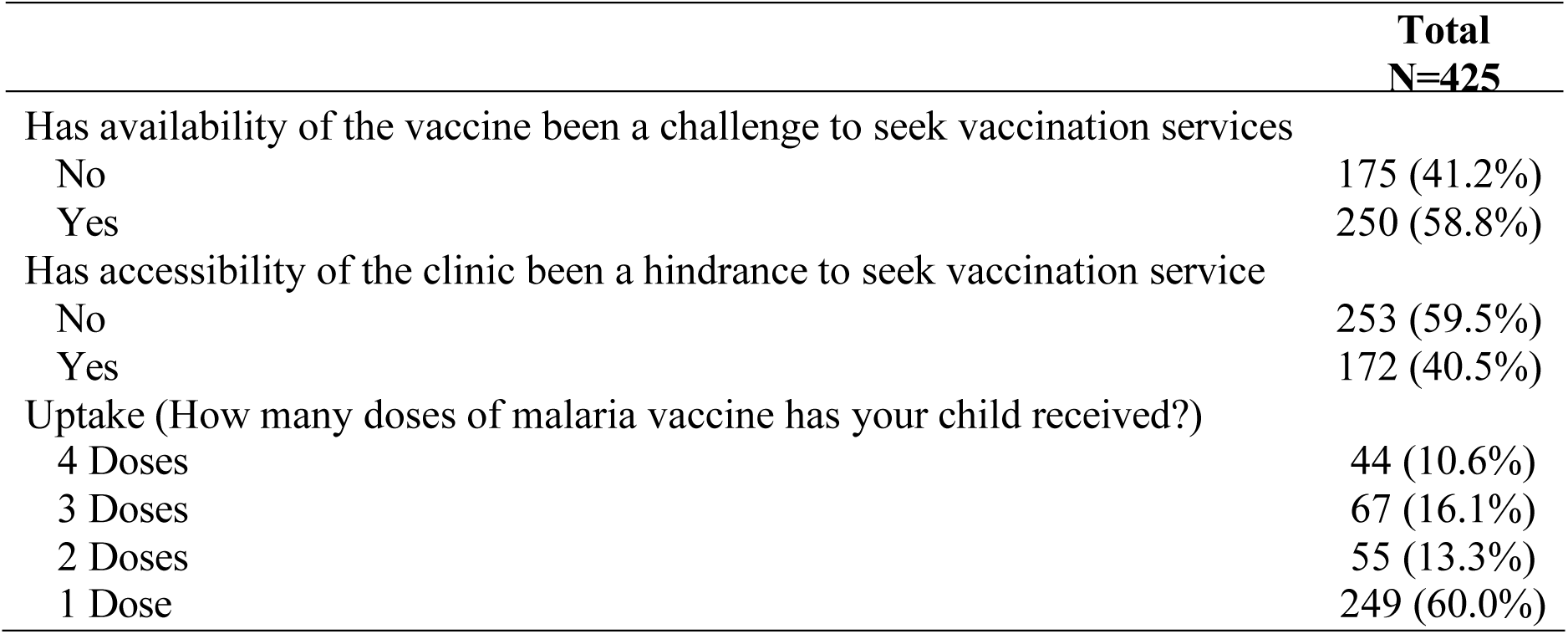
Health system factors associated with malaria vaccination services in Ugenya Subcounty, Siaya County.

### 4.3 Health System Factors Associated with Malaria Vaccination Services

#### 4.3.1 Health system factors associated with malaria vaccination services in Ugenya Subcounty, Siaya County

The study explored health system related factors that may be associated with malaria vaccine hesitancy. Access to health facilities, availability of malaria vaccines and malaria vaccine uptake were investigated.

Access to health facilities, availability of malaria vaccines and malaria vaccine uptake were investigated. A considerable proportion of caregivers 59.5% (n = 253) cited difficulty accessing clinics for vaccination services. This barrier was consistent with a report by KI, that malaria vaccine uptake was low due logistical challenges such as long distances to the health facilities and caregivers’ inability to afford transportation. As the informant stated in the following quote:

“You’re asking why vaccine uptake is poor, we have so many issues among them distance. Whereby, the distance from home to the facility will affect the mother. So, you also get transport. The mother will tell you that I didn’t have transport, that is why they didn’t come for the vaccines.” **KII02_MVH_NP**

Among the caregivers, 58.8% (n = 250) reported that the availability of the malaria vaccine was a concern. This finding was corroborated by a CHP who revealed that the vaccine was often unavailable at health facilities as quoted:

“Yes, it’s true they’re not available. I have a client who has gone to different facilities and has missed the vaccine.” **KII08_MVH_CHP.**

The lack of consistent supply had discouraged caregivers from returning for subsequent visits. This concern was expressed by a key informant in the following quote:

*“Yes. I have witnessed that happening. I have referred clients even three times they don’t get and this discourages them and when they finally get the vaccine, it is too late, that is the child is past the vaccine eligibility age.” **KII08_MVH_CHP***

Contrary to the above, the qualitative analysis revealed that stock outs were not common. One of the nurses stated that:

“On rare occasions, we have stock out for malaria vaccine. Though, since I’ve been here for a long time, I’ve never seen stock out for the vaccine. I have never seen a complete stock out. Okay. The quantity can be reduced, but I’ve never seen it.” **KII03_MVH_NP.** Further, a county health official reported that, “I’m not sure if we’ve had stock-outs. I doubt if we’ve had a stock-out of the vaccine.” **KII06_MVH_CHO.**

Sequential administration of the malaria vaccine showed a downwards trend. Ninety seven percent (n=415) of caregivers reported that their children had been vaccinated against malaria. Uptake of malaria vaccine revealed that 60% (n=249) of children child had received the first dose. However, the uptake of the subsequent doses dropped as 13.3% (n=55),16.1% (n=67) and 10.6% (n=44) of children had received the second, third and fourth doses of malaria vaccines, respectively (**Table 6)**. This was supported by a nurse practitioner as quoted.

**Table 6:** Bivariate analysis of health system factors associated with malaria vaccine hesitancy.

|  | No<br>N=244 | Yes<br>N=181 | p-value |
| --- | --- | --- | --- |
| Malaria vaccine availability |  |  |  |
| No | 93 (53%) | 82 (47%) | 0.14 |
| Yes | 151 (60%) | 99 (40%) |  |
| Accessibility of the clinic as a challenge |  |  |  |
| No | 185 (73%) | 68 (27%) | <0.001 |
| Yes | 59 (34%) | 113 (66%) |  |

“So, you get a child who has received the first two malaria vaccines. But on the second and third, there will be an issue. Because they are like, my kid has received MMR one and clinic is done. So, the only thing I’m waiting for is to come for weight and maybe deworming. So, they see that that’s not a big deal. Yeah. The main thing is the measles.” **KII02_MVH_NP**

#### 4.3.2 Association between health system factors and malaria vaccine

Two health system factors were analyzed for their association with malaria vaccine hesitancy among caregivers of children 6-59 months old (n= 425), at p ≤ 0.20 significance level.

The association between malaria vaccine availability and malaria vaccine hesitancy was significant (p = 0.14). Caregivers who indicated that malaria vaccine availability was not a challenge reported 47% (n=82) malaria vaccine hesitancy while those who indicated that it was a challenge reported 40% (n = 99) malaria vaccine hesitancy. Additionally, there was an association between clinic accessibility and malaria vaccine hesitancy (p < 0.001). Among caregivers who reported no difficulties accessing the clinic, 27% (n=68) were hesitant to malaria vaccine, compared to 66% (n=113) among those who faced accessibility challenges.

Qualitative analysis revealed that funding, leadership and governance, Health information system, Vaccine Supply Chain in relation to health system factors could have contributed to malaria vaccine hesitancy. The study also explored broader challenges within the Health System that could have affected Malaria Vaccine uptake, such as perception of vaccine effectiveness and systemic barriers to access.

#### 4.3.3 Funding of Malaria Vaccines

The study sought to find out the sources of financing for the malaria vaccines. The respondents reported that the provision and availability of the malaria vaccines were largely dependent on National Government support, which procured and distributed the malaria vaccines due to associated high cost This was illustrated in the following quote:

“Vaccines are the government that procures and distributes them.” **KII03_MVH_NP.** This was associated with the high cost of the vaccines as stated by a respondent “Everywhere it’s the government, vaccines are very expensive.” **KII03_MVH_NP.**

“Even if there is, I don’t have any idea, all I know is that the government supplies vaccines.” **KII02_MVH_NP**

#### 4.3.4 Leadership and Governance

In terms of malaria vaccination program, the study examined its leadership and governance. Respondents described the organizational framework of the programme within the health facilities. Specifically, they noted that the programme was led by the Kenya Expanded Programme on Immunization (KEPI) coordinator, who provided overall oversight and strategic direction. This leadership structure cascaded down to the nurse in charge, followed by the deputy in charge, and subsequently to other healthcare staff involved in malaria vaccine delivery. The hierarchical model of governance was illustrated by one respondent who stated:

“We have a KEPI coordinator, then a nurse in charge followed by the deputy in charge. Then other staff.” **KII03_MVH_NP**

#### 4.3.5 Health Education

Malaria vaccine education followed structured and community-based approach hich integrated both formal health systems and informal local systems. The process was typically initiated by nurses at health facilities, who provided health education to patients during visits to the health facilities. This information was then relayed to CHPs, chiefs, assistant chiefs, and village elders who served as vital intermediaries between the health system and the broader community. Village elders played a role in disseminating information through public forums such as barazas and the Nyumba Kumi initiatives. In addition, healthcare providers conducted outreach activities to trace and engage caregivers who had defaulted on vaccination schedules, reaching them directly in their homes or within local gathering points. These practices are supported by the quotes below:

“Health education. Where community dialogue is done at the community and when they come to the facility, we do health education.” **KII03_MVH_NP**

*“As a CHP one of my roles is to spread the news about malaria vaccine. So I mobilize other CHPs to share information with the rest of the community members. We liase with community get keepers now spreads the news through the barazas and nyumba kumi iniatives”*

**KII05_MVH_CHP**. Another CPH further mentioned that; “So you will find that different CHP are scheduled for outreaches in different locations within the same area, this helps cover for missed opportunities.” **KII08_MVH_CHP**

“After we have realized we have defaulters, we check the permanent register to assess those who have missed, where they are coming from and we organize for outreaches.” **KII03_MVH_NP** “Yeah. But at times we also do community outreach. And we carry the vaccines along with us” **KII02_MVH_NP**

#### 4.3.6 Supply Chain of Malaria Vaccines

Respondents reported that the vaccine delivery and storage process followed a carefully coordinated system to ensure the malaria vaccine remained potent and safe for use. The County KEPI coordinator took the lead by ordering a three-month stock from the regional depot in Kisumu, which was then stored in sub-county fridges. Nurses at local health facilities would make monthly orders and store the vaccines in designated cold storage rooms equipped with KEPI approved fridges. To preserve vaccine efficacy, they reported that a strict cold chain was maintained throughout. Vaccines were transported using cooler boxes fitted with dial thermometers to ensure temperatures stay between +2°C and +8°C. They highlighted that use of ice packs and proper vaccine carriers with shoulder straps were standard practice, and deliveries were made within 2–3 hours to avoid compromising potency. The process was handled with great care, avoiding rough transportation methods like placing vials on the backs of motorcycles, which could cause breakages due to poor road conditions as supported by below quotations;

“KEPI coordinator gets a three-month stock vaccines from Kisumu depot, which is the regional store. From a regional store to a KEPI store here in Ugenya sub-county. Then, from there, we order monthly and keep in our fridge. We have a specific room and specific fridge for storage. Because we must maintain the cold chain” **KII03_MVH_NP**

“When transporting we place dial thermometers inside the cooler box to check and maintain potency at temperatures of +2-+8, the minimum being +2 and maximum being +8. Two, being the minimum, maximum being positive eight degrees Celsius.” **KII02_MVH_NP.** Further the driver mentioned that; “We transport the vaccines using motorcycles and cooler boxes. Normally we are advised to deliver the vaccines within 2-3 hours for it to maintain their potency and to maintain a cold chain we use ice. During transportation, to ensure that the vaccines reach other facilities in good shape, we are forced to strap the carriers across our shoulders to prevent breakage of vaccine vials as they are made of glass”**KII01_MVH_DRV**

#### 4.3.7 Health System Challenges

Supply chain constraints, staffing shortages and unavailability of healthcare providers emerged as key barriers to effective delivery of the malaria vaccine in Ugenya Sub-County. Both health officials and community health promoters (CHPs) reported frequent vaccine stockouts, particularly during periods of increased demand. These stockouts limited the capacity of health facilities to consistently meet the needs of the children.

Logistical challenges further hindered distribution efforts. Poor road infrastructure – characterized by rough and often impassable terrain-posed significant risks during vaccine transportation. Respondents noted that transporting fragile glass vials on motorcycles along bumpy roads increased the likelihood of breakage, compromising both safety and vaccine supply.

These logistical setbacks were exacerbated by chronic understaffing at health facilities. In some instances, respondents described situations where caregivers were mobilized for vaccination only to find a single health care worker – or none at all-available at the facility. Such experiences discouraged caregivers from returning, particularly when they encountered closed or unattended clinics despite previous outreach efforts. These findings were highlighted in the following quotes: *“So, many times when there’s a rush of more people coming for it, many facilities experience stockouts.”* **KII05_MVH_CHO**

“Challenges that we experience are like poor road infrastructure. Being that the roads are bad, we can’t load at the back of the motorcycles as bumpiness of the road may cause breakage of vaccine vials as they are made of glass.” **KII01_MVH_DRV**

“Yes, there are few nurses who are not enough” **KII01_MVH_CHP**. Another CHP reported supported this by saying “The major challenges at times are that there is only one HCW in the facility and you have mobilized people to seek services at the HCF or at times the HCW are not available. This discourages some people, if you go and tell them to go, they will tell you that they went and didn’t find anyone at the facility, this discourages them.” **KII10_MVH_CHP**

#### 4.3.8 Compliance with Malaria Vaccine Schedule

Figure 6 depicts fluctuations in malaria vaccine uptake across the vaccination schedule highlighted the study.

**Fig 1:**
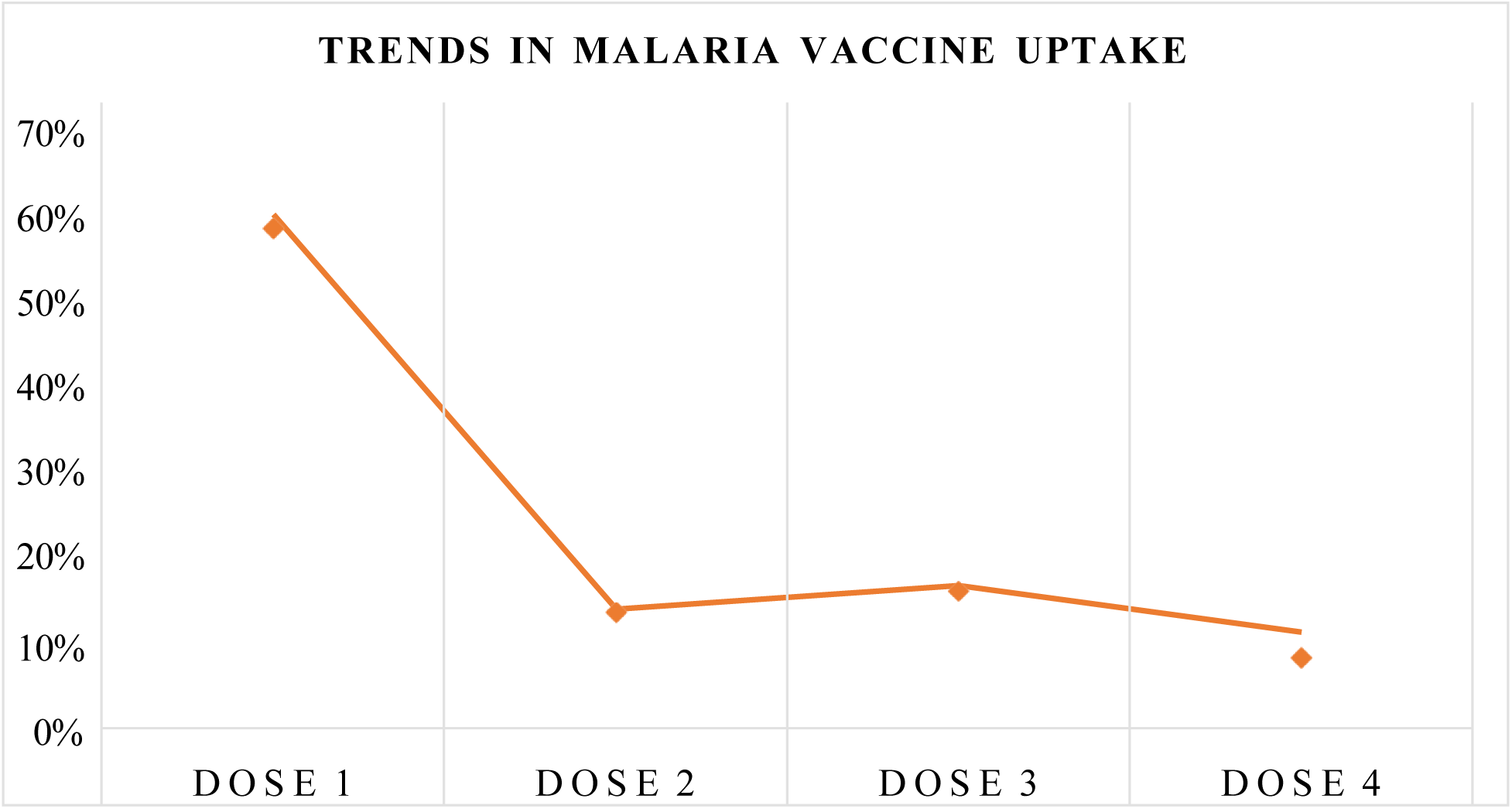
Malaria vaccine uptake trends among caregivers of 6-59 months children in Ugenya subcounty.

Healthcare providers and county health officials identified low uptake of the third and fourth malaria vaccine doses as major challenges affecting the overall success of the malaria vaccination programme While the uptake of the first and second doses were acceptable, there was a noticeable decline in the uptake of the third and fourth malaria vaccines doses, with later having the lowest uptake rate. This was supported by below quotes:

“…but there is a slight decrease from the first dose going to others, as you move to the other dose. January 2023 not 2024. But it keeps on fluctuating.” **KII06_MVH_CHO**. He furthers explains that: “But now the completion is not going well. But you know, the reason why maybe the first doses are doing well is because most of them are within the KEPI.” **KII06_MVH_CHO**. Another CHO corroborated the statement by stating that “So, the uptake basically you find from dose one, dose two up to three, the uptake is fairly okay. The challenge is the uptake of dose four. “Yeah, the uptake for dose one is okay. Let me just check what I’m talking about. But the uptake for dose one is okay. But dose four. Dose four generally is not, the uptake is not that good.”” **KII05_MVH_CHO, KII06_MVH_CHO**

Several underlying factors could have to contribute to this pattern. Healthcare workers reported that the long interval between the third and fourth doses often led to reduced follow-through, as caregivers lost motivation or simply forget the vaccine schedule over time. Additionally, there awas widespread misconception that childhood immunizations concluded at nine months of age. As a result, many caregivers stopped attending immunization clinics prematurely, before completing the full malaria vaccine schedule.

“And this is basically because of the intervals, the period in between dose three and dose four Is long.” **KII05_MVH_CHO**

*“Another thing that I’ve come to see. Most people tend to think vaccination ends at nine months. And before even we go to that, when you say they are reluctant, they are reluctant in that they are defaulters, or they don’t come for the vaccine at all? They are defaulters.”.” **KII02_MVH_NP**.* Another nurse fall.er reported that “They start, but they don’t finish.” **KII03_MVH_NP**

Despite relatively high levels of awareness and knowledge about malaria vaccine, these behavioral and systemic barriers could have contributed to low completion rates. In some sub-counties the fourth dose completion rate was below 50%. This was emphasized by the respondent in the following quote

“The completion is a problem. If it is the completion that is a true picture that we are also struggling even with now. I believe up to now, even our completion at dose four in most of the sub-counties barely less than 50%, so that gives us the information that now the community, even though they understand Malaria vaccine exists, they have not known the importance of completing the full dose for the vaccines that are available.” **KII05_MVH_CHO**

#### 4.3.9 Effectiveness of Malaria Vaccines Rollout

Malaria vaccine rollout in Ugenya Sub-County was a multifaceted process that incorporated both formal health systems mechanisms and practical field-level observations. Respondents highlighted the critical role of feedback from both community members and healthcare workers in evaluating the effectiveness of malaria vaccine implementation strategies. The participatory feedback mechanism was used to assess whether the strategies were being well executed as intended and identified areas for improvement. Confidence in the vaccine’s performance was further reinforced by evidence generated from previous trials, as well as ongoing surveillance through routine health records. As one respondent noted:

“But for us to have confidence that it is effective, then we rely mostly on findings from vaccine trials.” **KII06_MVH_CHO**

Uptake levels were tracked against target population estimates to provide insights into coverage rates and community response. Respondents also highlighted the importance of monitoring adverse events following immunizations (AEFI) as a key component of ongoing safety surveillance.

“And apart from that, again, you know, we also observe the adverse effects following immunization. So, in case you start implementing it and then we have those advance effects, those that are reported, those are also areas that are of our concern always. **KII05_MVH_CHO**

Routine programmatic activities – including supervisory visits, data review meetings, and monthly reporting were used to track progress, identify challenges and guide decision making as supported by the following quote:

“…and we get feedback on the effectiveness of the rollout strategies that are put in place, through supervision and review meetings, I believe we get that. And, even from the reports that we get on a monthly basis. So, if it is a specific antigen, you want to see how the uptake is because we already have a target population for the eligible and then you want to measure how the uptake is.”.” **KII05_MVH_CHO**

At the community level, local leaders in collaboration with CHPs played a important role of identifying households for sampling (for trials) and supported the administration of interviews to assess awareness, acceptance, and reasons for vaccine defaulting. Although formal evaluation tools were limited in scope, respondents expressed a general perception of transparency and accountability within the malaria vaccination program. The confidence appeared to be grounded in the assumption that the malaria vaccine performed comparably to other vaccines already integrated in the routine immunization schedule. This sentiment was echoed in the following quotes:

“Generally, we assume, just an assumption that people will take vaccines and the healthcare workers will be accountable, just like any other vaccines. Because we’ve been giving the other vaccines and we’ve not had any issues, we also assume that the same will apply to malaria vaccines.”” **KII06_MVH_CHO**

### 4.4 Predictors of malaria vaccine hesitancy among caregivers of children 6-59 months in Ugenya subcounty, Siaya County

The ten explanatory variables namely, *caregiver’s age*, *relationship with child*, *family size*, *marital status*, *household income*, *education level, awareness*, *knowledge, malaria vaccine availability* and *accessibility to the health facility* were found to be significantly associated with malaria vaccine hesitancy in the bivariate analysis (p ≤ 0.2).

These variables were included in a multivariable mixed-effects logistic regression model to identify the independent predictors of malaria vaccine hesitancy (**Table 8**). In the multivariable model, **caregiver’s age, marital status, family size, education level, knowledge and facility accessibility** remained significantly associated with malaria vaccine hesitancy.

Compared to caregivers aged 15–25 years, those aged 26–35 years had significantly higher odds (**2.59 times higher)** of reporting vaccine hesitancy (OR = 2.59, 95% CI [1.23 – 5.43], p = 0.012). Caregivers from household with family size of >=4 were significantly less likely to report malaria vaccine hesitancy (OR = 0.47, 95% CI [0.27 – 0.82], p = 0.008) compared to those with family size of <4, after controlling for other factors in the model.

Education level was statistically associated with malaria vaccine hesitancy. Caregivers with primary education had significantly lower odds of malaria vaccine hesitancy compared to those with no formal education (OR = 0.36, 95% CI [0.15–0.86], *p* = 0.021), while those with tertiary education had even lower odds (OR = 0.17, 95% CI [0.05–0.57], *p* = 0.004). Secondary education showed borderline significance in reducing hesitancy (OR = 0.39, 95% CI [0.15–1.01], *p* = 0.052). Caregivers with good knowledge of the malaria vaccine had borderline higher odds of malaria vaccine hesitancy compared to those with poor knowledge (OR = 3.13, 95% CI [1.00 – 9.76], p = 0.050).

Accessibility to health facilities was significantly associated with malaria vaccine hesitancy. Caregivers who reported challenges in accessing clinics were more likely to report hesitancy towards the malaria vaccine (OR = 4.82, 95% CI [2.96, 7.86], p < .001). Specifically, single caregivers had significantly higher odds of being hesitant compared to their married and widowed or separated counterparts (OR = 2.22, 95% CI [1.04–4.71], p = 0.039) and (OR = 0.61, 95% CI [0.23–1.63], p = 0.327) respectively.

**Table 7:** Multivariable analysis of factors associated with malaria vaccine hesitancy among caregivers of children 6-59 months in Ugenya subcounty Siaya County.

| Vaccine hesitancy | Odds ratio | P-value | [95% conf. | interval] |
| --- | --- | --- | --- | --- |
| **Age** |  |  |  |  |
| 15-25 | Ref |  |  |  |
| 26-35 | 1.30 | 0.442 | 0.67 | 2.54 |
| 36-45 | 2.59 | **0.012** | 1.23 | 5.43 |
| 45+ | 2.22 | 0.144 | 0.76 | 6.48 |
| **Relationship with child** |  |  |  |  |
| Biological parent | Ref |  |  |  |
| Guardian | 0.57 | 0.281 | 0.21 | 1.58 |
| **Family size** |  |  |  |  |
| < 4 | Ref |  |  |  |
| ≥ 4 | 0.47 | **0.008** | 0.27 | 0.82 |
| **Marital status** |  |  |  |  |
| Married | Ref |  |  |  |
| Separated/Widowed | 0.61 | 0.327 | 0.23 | 1.63 |
| Single | 2.22 | **0.039** | 1.04 | 4.71 |
| **Household income** |  |  |  |  |
| Below Ksh10000 | Ref |  |  |  |
| Ksh10000-15000 | 1.52 | 0.197 | 0.80 | 2.88 |
| Ksh15000+ | 1.16 | 0.711 | 0.52 | 2.60 |
| **Education level** |  |  |  |  |
| No formal Education | Ref |  |  |  |
| Primary Education | 0.36 | **0.021** | 0.15 | 0.86 |
| Secondary School Education | 0.39 | **0.052** | 0.15 | 1.01 |
| Tertiary Education | 0.17 | **0.004** | 0.05 | 0.57 |
| **Awareness** |  |  |  |  |
| No | Ref |  |  |  |
| Yes | 8.72 | 0.073 | 0.82 | 93.20 |
| **Knowledge** |  |  |  |  |
| Poor | Ref |  |  |  |
| Good | 3.13 | **0.05** | 1.00 | 9.76 |
| **Malaria vaccine availability** |  |  |  |  |
| No | Ref |  |  |  |
| Yes | 0.79 | 0.417 | 0.46 | 1.39 |
| **Clinic accessibility** |  |  |  |  |
| No | Ref |  |  |  |
| Yes | 4.82 | **<0.001** | 2.96 | 7.86 |

## CHAPTER FIVE: DISCUSSION

### 5.0 Overview of Vaccine Hesitancy

Malaria remains a significant public health concern with global efforts underway to eliminate the disease by 2030 (PATH, 2023). One of the recent developments in malaria control is the introduction of the malaria vaccine for children under the age of five, as a complementary intervention alongside existing prevention strategies. While vaccination is not the sole method for managing infectious diseases, it is the most effective strategy for their control, (Andre *et al*., 2008). Although the majority of individuals adhere to the recommended vaccination schedule, the effectiveness of immunization efforts is undermined by those who refuse or delay vaccinations. This study revealed that 42.9% (N=181)of caregivers had hesitancy towards the malaria vaccine.

According to the WHO, vaccine hesitancy is one of the top ten threats to global health and is driven by factors such as complacency, inconvenience in accessing vaccines, lack of confidence (WHO, 2019) among other factors. The high level of hesitancy observed in this study aligns with findings from other sub-Saharan African settings where vaccine coverage is sub-optimal (Larson *et al*., 2014). In 2022, a study conducted in Nigeria revealed that approximately 40.3% of caregivers reported childhood vaccine hesitancy, (Olaniyan *et al*., 2022). Similar trends of decline of malaria vaccine were observed in Ghana in 2021, (Yawson *et al*., 2021). Understanding factors associated with malaria vaccine hesitancy is critical in guiding policy makers, health workers, and social mobilizers on strategies to intensify efforts to promote malaria vaccine uptake and support the malaria eradication initiative in the Kenya to achieve the 2030 goal of Zero Malaria.

### 5.1 Socio-demographics and economic factors associated with malaria vaccine hesitancy among caregivers of children 0-59 months in Ugenya sub-County, Siaya County

*Caregiver’s age, marital status, family size* and *education level,* were significantly associated with malaria vaccine hesitancy. Caregivers between the ages of 36 – 45 were more likely to be malaria vaccine hesitant compared to the other age groups. Compared to younger caregivers, this age group often has greater authority in household health decisions. With more autonomy, they may question new interventions more critically, especially when weighing perceived risks versus benefits for their children, (Ojakaa *et al*., 2014). his finding concurs with that of Rajshekjar and colleagues who used machine learning in a longitudinal study to identify distinct population subgroups and their vaccine behaviors. The reported that middle-aged caregivers often approached vaccines with greater caution resulting in vaccine hesitancy (Rajshekhar et al. 2021). They suggested that this age group was likely exhibit vaccine hesitancy stemming from heightened awareness of potential side effects as well as stronger inclination to research and critical evaluation of health information before making decisions. Many individuals in this age group are also more active on social media, where they may be more exposed to misinformation, particularly surrounding newer vaccines like the one for malaria, (Kabir Sulaiman *et al*., 2023).

Additionally, people in this age group often juggle the responsibilities of caring for both children and aging parents. This dual caregiving role can increase the pressure to make safe and well-informed choices, contributing to their more cautious stance toward vaccination, (Achieng *et al*., 2020). An explorative qualitative study in Sierra Leone suggested that caregivers in this age group often have established beliefs and health practices shaped by years of experience, cultural norms, and community influence. If they have held negative perceptions about vaccines, experienced the vaccine side effects, or witnessed rumors and misinformation over time, they would be less receptive to newer vaccines, such as RTS,S for malaria, since it had not been included in the routine national vaccination schedule, (Jalloh *et al*., 2022)A cross-sectional study by Bugase and Tindana, (2024) in Ghana found no association between age and vaccine hesitancy. However, in the same study, mothers aged 35 years and above were more likely to accept the vaccines compared to those aged 20 years and below, possibly due to the previous experience with vaccines among the former group.

According to marital status, single caregivers were significantly more likely to report a higher level of malaria vaccine hesitancy compared to their married or widowed/separated counterparts. This is in concordance with a systematic review of immunization in sub-Saharan Africa which identified single motherhood as a notable barrier to full vaccination, often compounded by lack of or limited social support, constrained resources for clinic visits, and higher caregiving burdens, which affect healthcare prioritization and decision-making, including vaccination services, (Bangura *et al*., 2020) Married caregivers, on the other hand, were more likely to benefit from shared financial burden, joint decision-making and mutual encouragement regarding health-seeking behaviors, (Chinawa *et al*., 2024). In settings where healthcare decisions were influenced by family dynamics, Ozawa and colleagues suggested that the presence of a partner helped in reinforcing trust in health services and adherence to recommended vaccination schedules, (Ozawa *et al*., 2016). An exploratory cross-sectional mixed-method study conducted in the Kassena Nankana districts, Ghana, also reported that married mothers were likely to accept to vaccinate their children as compared to those not married due to the support, encouragement, and reminders given by their spouses and family members concerning vaccinations, (Bugase and Tindana, 2024).

Family size was also associated with malaria vaccine hesitancy. Caregivers from households with less than four members were significantly more likely to report malaria vaccine hesitancy compared to those from larger families. A systematic review of factors influencing childhood immunization uptake in Africa concur with the study finding by suggesting that this may be attributed to the fact that larger families tend to have more frequent interactions with the healthcare system (like antenatal clinic visits among others) for maternal and child healthcare, where they may get malaria vaccine information, which reinforces positive health behaviors. Additionally, they tend to have greater experience navigating child health services, having taken older siblings through routine immunization schedules successfully. This familiarity builds trust in vaccines and the health system over time, (Galadima *et al*., 2021). Further the review suggested that caregivers in larger households often prioritize preventive measures to avoid the higher cost and burden of treating illnesses that could easily spread among many children. As a result, they are likely to be more motivated to vaccinate all eligible children promptly, recognizing that prevention protects not just one child but the whole household’s well-being, (Galadima *et al*., 2021).

Previous research also suggested that larger family size could contribute to shared health knowledge and influence through social learning within households (Bbaale, 2013). A cross-sectional study in Angola indicated that families with more siblings had higher childhood vaccination uptake compared to smaller families. Our study findings align with the findings of a systematic review of the determinants of immunization in Africa, which identified family size as a factor influencing vaccine uptake. The review suggested that larger families often had strengthened social networks and shared decision-making, which in turn supported timely and complete immunization, (Bangura *et al*., 2020).

An inverse association was observed between education level and malaria vaccine hesitancy, suggested that caregivers with no formal education were more likely to be hesitant to those with some level of education. Most caregivers in the study had attained only primary education. Without clear, simple explanations, it would be hard for them to grasp how vaccines work or why completing all doses was important. This gap in understanding makes them more vulnerable to rumors, myths, or misleading stories they hear from people around them or on social media, (Adeyanju *et al*., 2022). This finding suggested the links among lower education levels and increased susceptibility to misinformation, limited vaccine confidence and poor health-seeking behavior (Hussain *et al*., 2019).

A study by Lin *et al*. (2020) reported that caregivers with higher education were better equipped to assess health risks and benefits, process medical information, and resist misinformation. Another study by Dubé and colleagues indicated that higher education correlates with increased vaccine acceptance (Dubé et al., 2013). According to Tadesse and colleagues, higher education lowered vaccine hesitancy by 60%, (Tadesse *et al*., 2022). This was likely due to better understanding of vaccine benefits, reduced susceptibility to misinformation, and improved health-seeking behavior. Qualitative analysis of our study suggested that poverty and illiteracy were among the factors that contributed to poor health choices. Given that malaria vaccine is relatively new and has a complex schedule, caregivers with lower levels of education would likely find it harder to understand the vaccination schedule and make informed decisions about the benefits compared to possible side effects.

Although the association between religion and malaria vaccine hesitancy was not statistically significant, analysis of qualitative findings suggested otherwise. Some religions did not embrace malaria vaccine. This pattern aligns with findings from a cross-sectional review of District Health Information System 2 (DHIS2) immunization data of Migori County in Kenya, by Shikuku et al., (2019), which reported that certain religious sects, including Legio Maria and Roho, discouraged vaccination altogether among its members, promoting the belief that healing occurred solely through spiritual means. These beliefs, often rooted in cultural and spiritual values, tend to spread through close-knit community networks, thus fueling distrust in modern medicine. Community-wide fears and spiritual doctrines often carry more weight than formal public health messaging. Our study findings align with the findings of a cross-regional analysis conducted across 195 regions that revealed that higher levels of religiosity and spirituality were consistently associated with increased vaccine skepticism, even when controlling for other demographic factors (Martens & Rutjens 2022).

In our study, there was no association between malaria vaccine hesitancy and other factors such as **sex of caregiver, caregiver’s relationship to child, household income, and employment status**. In concordance with our study, a multi-country cross-sectional study conducted in Democratic Republic of Congo, Senegal, Uganda and Nigeria found that sex of a caregiver did not affect vaccine hesitancy in community-based settings. The study was suggested that both men and women likely to share similar access, exposure, and beliefs about new vaccines thus making sex of the caregiver a less distinguishing factor, (Ndejjo *et al*., 2024). A household study in Guinea and Sierra Leone also found no association between caregivers’ sex and malaria vaccine hesitancy, especially in settings where both parents and family members were involved in making decisions on children’s health, (Ojakaa, 2014). Contrary to our findings, a systematic review by Bangura et al., (2020) revealed that male partners’ opposition was a more frequently cited barrier to childhood immunization. This study suggested that when fathers disapproved some healthcare services, they often withheld the social and financial support needed for female caregivers to complete their children’s immunization schedules, thus discouraging or preventing clinic visits altogether.

Lack of association between caregivers’ relationship to the child and malaria vaccine hesitancy was observed in our study. Aligning with our study, some published literature suggested that decisions about child vaccination were not shaped by the caregivers’ role in children’s lives but more by shared barriers such as fears of side effects, perceived vaccine safety, and the convenience of access to vaccination services. Additionally, the prevailing gender roles rather than parental status influenced the decision to vaccinate children. (Larson *et al*., 2014; Brown *et al*., 2022; Maamor *et al*., 2024; Jegede, 2007).

Our study found no significant association between caregivers’ employment status or household income and malaria vaccine hesitancy. This finding is echoed in a qualitative study by Essoh et al. (2023), which reported that household income had minimal influence on vaccine uptake among caregivers in Kenya. Supporting this, Favin *et al*. (2012) observed that even in low-income households, caregivers make concerted efforts to vaccinate their children when vaccines are accessible and perceived as necessary. Similarly, Oku et al. (2017) found that in many African settings, community-level beliefs and logistical challenges such as transport access and clinic proximity play a more prominent role in influencing vaccination behaviors than formal employment or income levels. These findings highlight the need to focus on social and structural enablers of vaccine uptake rather than solely targeting economic determinants.

On the other hand, some studies contradict this finding by suggesting that income can still play an indirect role. Rainey *et al*., (2011) showed that in settings where indirect costs like transport or lost work time are high, poorer households may struggle completing multi-dose vaccine schedules. Additionally, Méndez *et al*., study (2016) found that in low– and middle-income countries, wealth disparities predict immunization gaps due to hidden costs like travel expenses, time away from work, or informal payments, especially when health services are inconsistent. Taken together, these findings suggest that while free vaccination programs can reduce income-related barriers, complementary interventions such as addressing transport, service reliability, and community trust remain critical to achieving full coverage.

### 5.2 Individual and psychosocial factors associated with malaria vaccine hesitancy among caregivers of children 6-59 months in Ugenya sub county, Siaya County

Our study identified *knowledge* as significant factor influencing malaria vaccine hesitancy, although the association with knowledge was marginally significant. Majority of the caregivers had good knowledge about the vaccine, yet many still expressed hesitancies showing that awareness alone does not ensure acceptance or vaccination. This aligns with a study be Busby and colleagues, which suggested that trust in the source o f information and perceived risks played critical roles in vaccination decisions, (Busby *et al*., 2015). Our study findings revealed a gap between the knowledge about the malaria vaccine and completion the four doses of the vaccine.

Our study revealed that mainstream media and healthcare workers were primary sources of malaria vaccine information. Caregivers who cited healthcare workers and community media as source of information showed lower malaria vaccine hesitancy, echoing findings by Fine and colleagues that emphasized trust in healthcare workers as a strong determinant of vaccine acceptance (Fine *et al*., 2011). Conversely, Jamison and Colleagues suggested that people who relied on social media for information were susceptible to digital misinformation hence likely to be vaccine skeptics, (Jamison *et al*., 2020).

Our study did not find a significant link between with other psychosocial factors like willingness, awareness, perceived risk, and malaria vaccine hesitancy, however qualitative findings suggested that these factors played an important role in shaping caregiver decisions. Healthcare providers reported that some caregivers expressed worries about possible side effects of vaccines such as mild reactions and paralysis, which resulted in malaria vaccine hesitancy. These findings are consistent with those of a qualitative descriptive study conducted in Kenya which highlighted safety concerns of vaccine as a primary factor in vaccine hesitancy, with mothers expressing fears of adverse, (Kimotho, 2025; Larson et al. 2014) suggested that rumors and past adverse events real or perceived could amplify fear and fuel vaccine hesitancy, especially in communities with limited access to clear, trustworthy information.

A study by Jegede (2007) on polio vaccine refusal in northern Nigeria revealed that fears of serious side effects, including paralysis, were deeply rooted in local narratives and mistrust, which spread easily by word of mouth. These fears worsened when health workers failed to communicate openly about possible side effects, leaving space for misinformation to thrive. As Larson *et al*. (2014) noted, perceived risk was often rooted in emotional, social, and cultural factors rather than just factual knowledge. Even when individuals understood the benefits of vaccination, their actions would occasionally still be driven by fear especially if these fears are reinforced by past experiences, peers, or community-level misinformation.

A study by Busby *et al*., (2015) found that a general willingness to vaccinate did not always translate to consistent follow-through if underlying doubts remained unaddressed. Willingness is not a simple binary choice (yes/no) but exists along a continuum, shaped by social context and trust, (Piltch-Loeb & DiClemente, 2020). In the context of our study, willingness meant that the caregivers would allow their children to get the malaria vaccine. According to Dube et al., (2013), “vaccine decision-making is dynamic and socially negotiated, often influenced by peer experiences, word of mouth, and confidence in the healthcare system”. They seem to suggest that simply providing information about the vaccine was not enough to guarantee uptake. Instead, communication strategies should aim to build trust, address specific fears, and involve community voices to strengthen caregivers’ confidence in their choices. A supportive community environment where caregivers see positive examples could help convert hesitant willingness into concrete action, ultimately improving completion of multi-dose regimens like the RTS,S malaria vaccine.

### 5.3 Health system factors associated with malaria vaccine hesitancy among caregivers of children 6-59 months in Ugenya sub county, Siaya County

The study identified *access to the health facilities* as a critical determinant of malaria vaccine hesitancy, with significant differences observed between caregivers who reported access and those who did not. Notably, nearly half of the caregivers in this study cited distance to health facilities as a major barrier to malaria vaccine vaccination. In concurrence with the WHO’s global reports, geographic and logistical barriers such as long distances, lack of transportation, and poor health infrastructure are among the most common constraints to vaccine uptake in low-resource settings, (WHO, 2023). *Aaby et al.*, (2020) and Masters *et al*., (2017), also reported that physical access to health facilities not only influenced whether a child would be vaccinated but also parental perceptions of vaccine importance and timeliness. They suggested that in many rural or underserved areas such as our study settings, caregivers would delay or forgo vaccination due to the difficulties in reaching a facility especially when multiple doses were required over time, as is the case with the RTS,S malaria vaccine. Additionally, a study by Melendez-Torres *et al*. concluded that poor road conditions, long waiting times, and clinic hours that didn’t suit the caregivers’ schedules significantly reduced vaccine compliance in rural Kenya and Uganda (Melendez-Torres *et al*., 2021).

Even though availability of malaria vaccine was not significantly associated with malaria vaccine hesitancy, more than half of the caregivers in our study reported it as a challenge, with 58.8% indicating they had faced difficulties finding the vaccine when needed. However, this perception was not supported by some healthcare workers who reported that stockout were rare. This difference between caregivers’ perceptions and providers’ reports suggested a possible communication gap rather than a consistent supply chain problem. Therefore, when caregivers encountered temporary delays, scheduling mismatches, or lack of clear information about when the vaccine were offered, they were likely to misinterpret these as stock-outs thus undermining their confidence (Melendez-Torres *et al*., 2021; WHO, 2019).

Also, this disconnect between healthcare providers and caregivers on malaria vaccine availability may be partly explained by the structure of Kenya’s immunization program, where the national government acts as the sole supplier and funder of the RTS,S malaria vaccine. WHO report suggested that while centralized procurement, such as the Kenya immunization program, ensured consistent policy and supply chain oversight, it could also create bottlenecks when local distribution and communication were weak, (WHO, 2023). According to Cutts *et al*., (2013) and Melendez-Torres et al. (2021), even minor delays in distribution, lack of real-time information, or poor scheduling communication could create the perception of vaccine unavailability to the caregivers thus undermining their trust in the health system. Whilst vaccine supply may be stable nationally, local gaps in communication or transport logistics, such as late vaccine deliveries or distribution in smaller batches could fuel rumors of stockouts at community level.

Our study revealed a steady decline in in uptake across the required four doses of the RTS,S malaria vaccine, with the sharpest drop-off reported on the third and especially the fourth doses Major reason given for these trends was family relocation from a malaria endemic county where the malaria vaccine was offered to a county where it wasn’t offered. This mismatch between where the vaccine was offered and where families lived or moved highlighted a larger gap in Kenya’s malaria vaccination policy. This problem is not unique to Kenya. Gavi (2021), noted that inconsistent access and internal migration in low– and middle-income countries (LMICs) frequently led to incomplete immunization, undermining the collective goal of building herd immunity. Similarly, a World Health Organization (2021) report emphasized that families who relocated across regions without harmonized vaccination services were at high risk of missing scheduled doses, which limits the overall effectiveness of multi-dose vaccines.

In addition to geographic barriers, timing also played a role. According to the Health officials interviewed, it was pointed out that the wide gap between the third and fourth dose contributed to missed appointments, pointing to a knowledge-practice gap (Busby *et al*., 2015). The common belief that childhood immunization ended at nine months also contributed to lapses in completion of the malaria vaccine schedule. Moreover, the drop-off seen in the later doses may reflect what Larson et al. (2014) describe as vaccine fatigue, where the initial enthusiasm or sense of urgency fades when no immediate threat is visible. Caregivers may start with high motivation but become less vigilant about follow-up doses, especially when daily demands compete for their attention.

Our study found that perceptions of the effectiveness of malaria vaccine were shaped less by formal, data-driven evidence and more by trust in the health system and observed community experiences. Healthcare providers and officials cited community feedback, informal supervision reports, and routine records as key indicators of success The heavy reliance on anecdotal feedback raises valid concerns echoed by the World Health Organization and Gavi (2021), who warned that absence of rigorous monitoring can allow gaps in service delivery to go unnoticed, potentially weakening long-term vaccine confidence and effectiveness (Scobie *et al*., 2020: Gavi, 2021).

Despite broad awareness about malaria vaccination, some caregivers in the study still missed follow-up doses or disregarded referrals from CHPs. This finding suggested that community-level complacency may contribute to malaria vaccine hesitancy, a pattern consistent with the WHO’s 3C model (2015), which identifies complacency as a key determinant of vaccine hesitancy. In high-burden settings where malaria is endemic, the disease is often perceived as a routine or “normal” risk, reducing the perceived urgency of preventive interventions such as vaccination. This complacency is frequently reinforced by the influence of informal social networks, where guidance from neighbors, family members, or religious leaders may hold greater sway than information disseminated by healthcare professionals, thereby weakening the impact of formal health communication (Dube et al., 2013; Larson et al., 2014).

This finding of wide awareness with low malaria vaccine uptake highlights that fact that awareness alone does not guarantee vaccine completion. As research by Dube *et al*. (2013) and Ozawa *et al*. (2017) suggested, building trust requires deeper community engagement. Public health campaigns are more effective when they empower key and trusted community gatekeepers such as religious leaders, community elders, and CHPs who can translate health messages into culturally meaningful dialogue. Stronger collaboration between frontline health workers and community influencers could help address misinformation, reshape norms, and reduce hesitancy.

Our study’s findings affirmed the critical role that leadership and governance structures play in ensuring the effective delivery and acceptance of the malaria vaccine. The clear chain of command, stretching from KEPI coordinators down to nurses in charge, reflected strong alignment with the WHO’s Health System Building Blocks framework, which emphasizes that robust leadership enhances accountability, service quality, and consistency (WHO, 2007). In practice, this structure helps to maintain vaccine availability by supporting coordinated procurement, distribution, and cold chain management all of which serve to reduce the risk of stockouts that could fuel hesitancy if caregivers repeatedly found vaccines unavailable when they visit clinics.

Equally important is how health communication and community engagement are operationalized at the grassroots level. The involvement of Community Health Promoters (CHPs) and local gatekeepers in our study reflected effective use of trusted community structures to share accurate information, counter rumors, and encourage caregivers to complete all four doses of the RTS,S vaccine. This was consistent with the findings of Dube *et al*. (2013) and Larson et al. (2014), which highlighted that trusted intermediaries were vital in addressing concerns and misconceptions, particularly in settings where misinformation about side effects or new vaccines could quickly spread.

Despite the sound governance structure, our study found that vaccine hesitancy still emerged when availability was inconsistent. Reports of caregivers arriving at health centers only to find the malaria vaccine out of stock due to supply chain disruptions or miscommunication eroded trust in the system and discouraged follow-up visits. This aligns with Cutts *et al*., (2013) findings, that suggested that perceived unreliability of vaccine supply undermined caregivers’ confidence, even when vaccines were free and technically accessible. Altogether, strong leadership, clear hierarchies, and effective community engagement could ensure steady vaccine availability and build trust. However, without continuous supply and real-time communication about vaccine schedules, even the best-designed governance structures will likely fall short leaving gaps that fuel hesitancy.

The findings of our study revealed that while the cold chain and transportation of the malaria vaccine in rural areas appeared sound, the system were fragile and prone to disruptions that directly influenced vaccine hesitancy. Vaccine transporter in the study highlighted how the fragility of glass vials, poor road conditions, and occasional power outages threatened the integrity of vaccine storage, raising doubts about the vaccine’s safety and potency. Consistent with WHO (2022) guidelines, careful cold chain handling that used of dial thermometers and ice packs was critical to maintain vaccine efficacy instead. This requires dependable transport infrastructure that is rarely found in many rural facilities, similar to our study settings. Recurrent stockouts during periods of high demand, and staffing shortages, meant that caregivers would sometimes arrive at health centers only to find the vaccine unavailable, a scenario that not only wasted their time but also undermined their trust in the health system. Aaby *et al*., (2020) suggested that similar systemic gaps were likely to create fertile ground for rumors, fear and reluctance that fueled vaccine hesitancy even when caregivers were willing to get their children vaccinate.

## CONCLUSION

This study identified several socio-demographic, psychosocial, and health system factors significantly associated with malaria vaccine hesitancy among caregivers of children aged 6–59 months in Ugenya Sub-County, Siaya County. Caregivers aged 36–45 years were more likely to exhibit malaria vaccine hesitancy, reflecting heightened scrutiny of new health interventions, greater exposure to misinformation, and entrenched health beliefs. Single caregivers were also more hesitant to malaria vaccine than married caregivers, suggesting the importance of social and financial support in health decision-making. Larger households (≥4 members) were less hesitant, potentially due to increased familiarity with child health services and shared decision-making.

Education emerged as a protective factor, with higher levels of education strongly associated with lower malaria vaccine hesitancy, highlighting the vulnerability of low-literacy caregivers to misinformation. While most caregivers had good knowledge of the malaria vaccine, this did not translate into uptake, underscoring the role of mistrust and misinformation. Accessibility to health facilities was a critical determinant, with difficulty in reaching clinics markedly increasing hesitancy.

Qualitative findings revealed that certain religious beliefs, negative perceptions of vaccine side effects, and perceptions of stockouts influenced decision-making. Even when national vaccine supply was stable, logistical bottlenecks and poor communication created perceptions of unreliability, undermining trust in the health system.

The findings indicate that reducing malaria vaccine hesitancy requires a multifaceted approach that addresses trust, accessibility, and service reliability. Health communication must extend beyond information dissemination to actively counter misinformation through trusted community gatekeepers, religious leaders, and CHPs. Reliable vaccine availability is essential perceptions of stockouts, even if inaccurate, can rapidly erode caregiver confidence of malaria vaccine. Additionally, social support networks significantly shape vaccination decisions, necessitating targeted interventions for single caregivers. Geographic and logistical barriers, such as long travel distances and poor road conditions, can hinder completion of multi-dose regimens like RTS,S, especially with long intervals between doses. The centralization of vaccine supply at the national level calls for robust county-level distribution planning and transparent communication to prevent perceptions of inequitable or unreliable access. Strengthening formal monitoring while investing in local trust networks offers a promising pathway to high RTS,S uptake as Kenya moves toward national scale-up.

Addressing malaria vaccine hesitancy in Siaya County requires a comprehensive, community-centered approach that extends beyond the mere provision of the vaccines.

## Data Availability

Data will be available upon request

